# Preterm birth rates in a large tertiary Australian maternity centre during COVID-19 mitigation measures

**DOI:** 10.1101/2020.11.24.20237529

**Authors:** Linda A. Gallo, Tania F. Gallo, Danielle J. Borg, Karen M. Moritz, Vicki L. Clifton, Sailesh Kumar

## Abstract

**Objectives:** To compare the prevalence of live preterm birth rates during COVID-19 restriction measures with infants born during the same weeks in 2013-2019 in Queensland, Australia.

**Design, setting, participants:** Deidentified obstetric and neonatal data were extracted from the Mater Mothers’ electronic healthcare records database. This is a supra-regional tertiary perinatal centre.

**Main outcome measures:** Logistic regressions were used to examine preterm birth rates during the beginning of COVID-19 restrictions (16 March-17 April; “early”; 6,955 births) and during the strictest part of COVID-19 restrictions (30 March-1 May; “late”; 6,953 births), according to gestational age subgroups and birth onset (planned or spontaneous). We adjusted for multiple covariates, including maternal age, body mass index, ethnicity, parity, socioeconomic status, maternal asthma, diabetes mellitus and/or hypertensive disorder. Stillbirth rates were also examined (16 March-1 May).

**Results:** A reduction in planned moderate/late preterm births was observed primarily during the early restriction period compared with the same calendar weeks in the previous seven years (29 versus an average of 64 per 1,000 births; adjusted odds ratio [aOR] 0.39, 95% CI 0.22-0.71). There was no effect on extremely or very preterm infants, spontaneous preterm births, or stillbirth rates. Rolling averages from January to June revealed a two-week non-significant spike in spontaneous preterm births from late-April to early-May, 2020.

**Conclusions:** Planned births for moderate/late preterm infants more than halved during early COVID-19 mitigation measures. Together with evidence from other nations, the COVID-19 pandemic provides a unique opportunity to identify causal and preventative factors for preterm birth.

## Introduction

The 2020 novel coronavirus (COVID-19) (1) was declared a pandemic by the World Health Organization on March 11^th^, 2020. In Australia, preventative measures, including staying at home if feeling unwell, social distancing, self-isolation for international arrivals, and cancellation of large gatherings were instituted from mid-March (2). In Queensland specifically, all but essential services were shut down by the end of March, with gradual easing of restrictions commencing in early May.

From a global perspective, the pandemic has resulted in significant reconfiguration and reduction of routine healthcare services. Often, only life-saving and emergency procedures were available with cessation of routine screening programs and elective surgery. Unfortunately, this strategy has resulted in significant unintended consequences – delay in cancer diagnosis, poor control of chronic cardiovascular and metabolic disorders, and profound psychological stress amongst others. In maternity care however, consequences of the imposed restrictions have been mixed. Early reports from some countries suggest that although stillbirth rates increased (3-5), rates of preterm birth actually decreased (5-9). It is not clear if there is a similar trend in Australia.

The aim of this study was thus to investigate preterm birth rates and trends and stillbirths during the lockdown period in a single tertiary perinatal centre in Queensland.

## Materials and Methods

This study was performed at the Mater Mothers’ Hospital in Brisbane. This is a supra-regional tertiary perinatal centre with ∼10,000 births per year. Ethical and governance approvals were obtained from the institution’s Human Research Ethics Committee and Governance and Privacy office respectively (Ref No: HREC/MML/61799). We analysed preterm birth rates during the strictest period of lockdown (30 March-1 May; “late”). To examine effects of earlier mitigation measures, we also analysed an equal 33-day period commencing two-weeks earlier (16 March-17 April; “early”). Each period was compared with the exact same calendar period for seven years (2013-2019) preceding the COVID-19 pandemic. We assessed stillbirth rates during the entire study period (16 March-1 May).

### Study design and participants

Deidentified obstetric and neonatal data were extracted from the hospital’s electronic healthcare records database. Women with singleton pregnancies who birthed between 16^th^ March to 1^st^ May in years 2013-2020 were included in the main analyses. Preterm births were categorised as follows: 23+0–27+6 weeks (extremely preterm); 28+0–31+6 weeks (very preterm); and 32+0–36+6 weeks (moderate/late preterm) and only livebirths were included. The comparison group was live infants born at term (≥37 weeks’ gestation). Preterm birth rates were also analysed according to birth onset: planned (Caesarean section or induction of labour) or spontaneous. Fourteen-day rolling averages (7 days prior to 6 days after) were calculated for the percentage of preterm births from January to June each year. To eliminate effects of whole-year shifts in preterm birth prevalence, rolling averages were also presented as a percentage change from the average preterm birth for that year. For singleton stillbirths reported, all gestational ages were included.

In supplementary analyses, preterm birth rates for women with multiple pregnancy (twins and triplets) who birthed between 16^th^ March to 1^st^ May from 2013-2020 were similarly calculated. The comparison group was multiple infant sets born at term (≥37 weeks’ gestation) where at least one infant in the set was born live.

### Statistical analyses

Logistic regressions were performed for each restriction period to compare the probability of preterm versus full-term births in 2020 compared with consolidated 2013-2019 data. This was also performed for planned and spontaneous births separately. Multinomial logistic regressions were performed to assess the odds of being born in each preterm category versus the reference full-term category in 2020 compared with consolidated 2013-2019 data. The odds of preterm birth in each preceding year was also compared using logistic regressions, with year 2020 set as the reference year. Covariates in the adjusted models were key contributors to preterm birth, including maternal age, body mass index, ethnicity, parity, socioeconomic status, and history or current asthma, diabetes mellitus, and/or hypertensive disorder based on signal differences (*P*≤0.25) in the distribution of cases between years by ANOVA (scale) or Chi-square testing (categorical). Socioeconomic status was represented by tertiles of the Socioeconomic Indexes for Areas (SEIFA, 2016) index of relative socioeconomic advantage and disadvantage scores for the maternal postcode of residence (lower, middle, upper). Maternal smoking status around the time of conception and/or the first trimester of pregnancy were not included in the adjusted model as these did not differ between the years. Information pertaining to alcohol consumption around the time of conception and/or the first trimester of pregnancy was also not included in the adjusted model as data was available for only 50% of the population each year. The prevalence of stillbirths was compared between the years using Chi-square testing.

For multiple pregnancies, the odds of preterm versus full-term birth in each preceding year was compared with year 2020 using a logistic regression. Maternal ethnicity and socioeconomic status were included in the adjusted model based on signal differences (*P*≤0.25) in the distribution of cases between years by Chi-square testing.

## Results

In the last eight years from mid-January to mid-June, the proportion of all preterm births was at its lowest level at the end of March to mid-April, 2020, which coincides with the implementation of COVID-19 restrictions (Figure 1A, *N*=2,973 preterm infants/35,028 full term infants from Jan 8^th^-Jun 21^st^ 2013-2020). This was primarily attributed to a reduction in planned preterm births (Figure 1B, C). When adjusted for inter-year variability, preterm birth prevalence in 2020 remained low at the end of March to mid-April compared with all other years, primarily attributed to a decline in planned preterm births (Figure 2A, B, C). There was a two-week spike in spontaneous preterm births from the end of April to early May, 2020 (Figure 1C, 2C).

**Figure 1.**
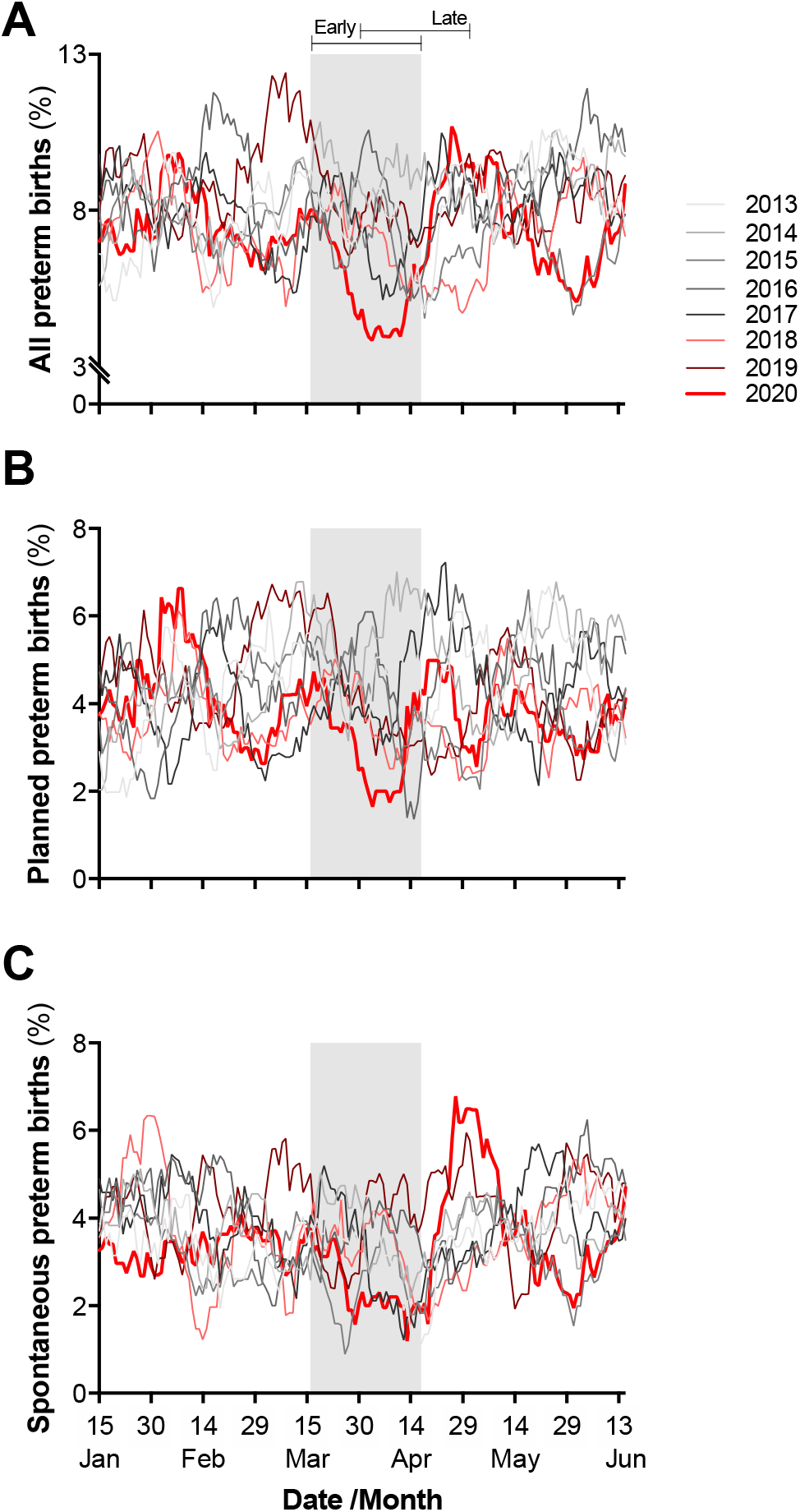
Proportion of preterm singleton livebirths. Proportion of A) all preterm births, B) planned preterm births (birth onset by Caesarean section or induction), and C) spontaneous preterm births. Rolling 14-day average from mid-January to mid-June in years 2013-2020. Plotted dates correspond to average from 7 days prior to 6 days after. Shading represents the two study periods, with dark grey indicating the overlapping study period. Rolling averages calculated from *N*=2,973 preterm infants/35,028 full term infants born on Jan 8^th^-Jun 21^st^, 2013-2020.

**Figure 2.**
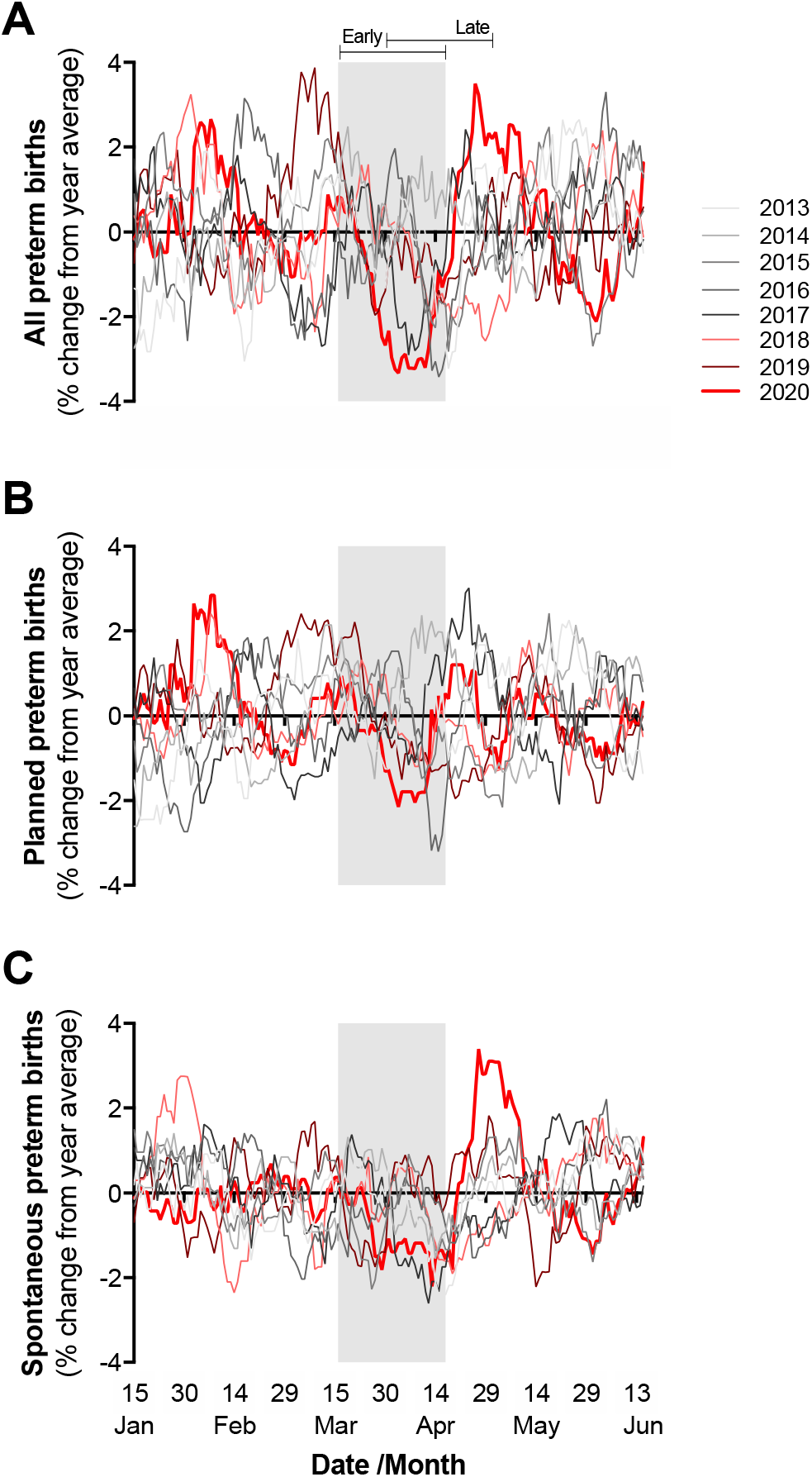
Percentage change in preterm singleton livebirths from yearly average. Proportion of A) all preterm births, B) planned preterm births (birth onset by Caesarean section or induction), and C) spontaneous preterm births normalised by subtracting the average preterm prevalence of January to June for each specific year. Rolling 14-day average from mid-January to mid-June in years 2013-2020. Plotted dates correspond to average from 7 days prior to 6 days after. Shading represents the two study periods, with dark grey indicating the overlapping study period. Rolling averages calculated from *N*=2,973 preterm infants/35,028 full term infants born on Jan 8^th^-Jun 21^st^, 2013-2020.

Table 1 shows maternal characteristics of all singleton livebirths between March 16 and May 1 in years 2013-2020. Subsequent analyses for the “early” period (March 16-April 17) included 6,955 births (510 preterm; 48 extremely preterm, 69 very preterm, 393 moderate/late preterm) and for the “late” restriction period (March 30-May 1), 6,953 births were included (501 preterm; 42 extremely preterm, 64 very preterm, 395 moderate/late preterm).

**Table 1.** Maternal characteristics of singleton livebirths between March 16-May 1 by year.

|  | 2013 | 2014 | 2015 | 2016 | 2017 | 2018 | 2019 | 2020 |
| --- | --- | --- | --- | --- | --- | --- | --- | --- |
| **Total N** | 1,194 | 1,262 | 1,269 | 1,254 | 1,233 | 1,244 | 1,299 | 1,231 |
| **Age [median, IQR, (N)]<sup>a</sup>** | 31.8, 6.6 (1,187) | 31.9, 6.7 (1,259) | 31.9, 6.7 (1,262) | 32.3, 6.9 (1,248) | 32.0, 6.6 (1,230) | 32.4, 6.5 (1,238) | 31.9, 6.8 (1,295) | 32.7, 6.8 (1,226) |
| **BMI [median, IQR, (N)]<sup>a</sup>** | 23.1, 6.3 (1,187) | 22.9, 6.3 (1,259) | 22.9, 6.0 (1,262) | 22.7, 5.5 (1,248) | 22.7, 5.6 (1,230) | 22.8, 6.2 (1,238) | 22.8, 5.5 (1,295) | 23.3, 6.1 (1,226) |
| **Ethnicity [%, (N)]<sup>a</sup>** |  |  |  |  |  |  |  |  |
| *Caucasian* | 64.9 (772/1,189) | 58.1 (733/1,261) | 63.1 (798/1,265) | 65.5 (821/1,254) | 64.7 (796/1,231) | 62.4 (776/1,244) | 61.2 (791/1,292) | 61.2 (752/1,229) |
| *Aboriginal and Torres Strait Islander* | 2.3 (27/1,189) | 2.9 (36/1,261) | 2.5 (31/1,265) | 2.0 (25/1,254) | 1.9 (23/1,231) | 2.6 (32/1,244) | 2.3 (30/1,292) | 2.0 (24/1,229) |
| *Asian* | 14.2 (169/1,189) | 14.8 (187/1,261) | 13.5 (171/1,265) | 14.1 (177/1,254) | 13.1 (161/1,231) | 14.7 (183/1,244) | 13.3 (172/1,292) | 13.9 (171/1,229) |
| *South Asian* | 6.9 (82/1,189) | 10.2 (129/1,261) | 7.7 (97/1,265) | 7.3 (92/1,254) | 8.9 (109/1,231) | 7.3 (91/1,244) | 10.0 (129/1,292) | 10.3 (127/1,229) |
| *Other* | 11.7 (139/1,189) | 14.0 (176/1,261) | 13.3 (168/1,265) | 11.1 (139/1,254) | 11.5 (142/1,231) | 13.0 (162/1,244) | 13.2 (170/1,292) | 12.6 (155/1,229) |
| **Parity [%, (N)]** |  |  |  |  |  |  |  |  |
| *0* | 26.5 (316) | 39.2 (495) | 50.0 (634) | 50.2 (629) | 66.5 (820) | 68.5 (852) | 72.7 (945) | 74.2 (911) |
| *1* | 48.5 (579) | 40.7 (514) | 31.0 (394) | 31.1 (390) | 20.6 (254) | 19.1 (238) | 15.9 (207) | 17.4 (214) |
| *2* | 17.4 (208) | 11.5 (145) | 11.7 (148) | 11.1 (139) | 6.9 (85) | 6.4 (79) | 6.9 (90) | 4.8 (59) |
| *3+* | 7.6 (91) | 8.6 (108) | 7.3 (93) | 7.7 (96) | 6.0 (74) | 6.0 (75) | 4.4 (57) | 3.6 (44) |
| **Residence socioeconomic status [%, (N)]** |  |  |  |  |  |  |  |  |
| *Low* | 14.8 (177) | 13.2 (167) | 14.7 (186) | 15.1 (189) | 14.0 (173) | 16.2 (202) | 14.7 (191) | 13.0 (160) |
| *Middle* | 12.6 (150) | 15.5 (196) | 18.4 (234) | 17.9 (224) | 16.8 (207) | 17.0 (212) | 15.9 (207) | 17.5 (216) |
| *Upper* | 72.6 (867) | 71.2 (899) | 66.9 (849) | 67.1 (841) | 69.2 (853) | 66.7 (830) | 69.4 (901) | 69.5 (855) |
| **History or current chronic disease [%, (N)]<sup>a</sup>** |  |  |  |  |  |  |  |  |
| *Asthma* | 18.9 (225/1,191) | 17.6 (222/1,261) | 17.2 (218/1,267) | 17.4 (218/1,253) | 17.0 (209/1,229) | 16.2 (201/1,243) | 15.6 (203/1,298) | 14.1 (174/1,231) |
| *Diabetes* | 6.7 (78/1,161) | 7.3 (90/1,228) | 8.4 (106/1,264) | 8.2 (102/1,239) | 8.9 (107/1,209) | 9.7 (119/1,222) | 8.0 (103/1,281) | 11.8 (143/1,207) |
| *Hypertensive disorders* | 7.8 (93/1,194) | 6.5 (82/1,262) | 5.9 (75/1,269) | 5.5 (69/1,254) | 5.7 (70/1,233) | 4.6 (57/1,244) | 4.7 (61/1,299) | 3.9 (48/1,231) |
| **Smoking [%, (N)]<sup>a,b</sup>** | 12.8 (152/1,187) | 12.0 (142/1,188) | 12.1 (145/1,199) | 11.6 (143/1,231) | 11.9 (145/1,217) | 10.9 (133/1,215) | 12.9 (164/1,268) | 11.1 (34/1,207) |
| **Alcohol consumption [%, (N)]<sup>a,b</sup>** | 38.4 (242/631) | 31.9 (200/626) | 35.5 (221/622) | 29.2 (177/607) | 30.6 (185/604) | 44.3 (273/616) | 41.4 (281/678) | 35.2 (225/639) |
| **Medically indicated birth onset [%, (N)]** |  |  |  |  |  |  |  |  |
| *Caesarean section* | 27.5 (328) | 23.9 (301) | 22.5 (286) | 22.2 (278) | 19.4 (239) | 18.9 (235) | 20.2 (262) | 20.8 (256) |
| *Induction of labour* | 28.4 (339) | 29.4 (371) | 32.5 (412) | 33.8 (424) | 37.0 (456) | 37.8 (470) | 37.4 (486) | 41.1 (506) |

<sup>a</sup> Some missing data for age, body mass index, ethnicity, chronic disease, smoking and alcohol consumption, with total *N* indicated in parentheses, <sup>b</sup> Smoking and alcohol consumption in early pregnancy.

During the “early” period in 2020, 49 per 1,000 singleton livebirths were preterm versus an average of 77 during the same calendar weeks of the previous seven years (aOR 0.62, 95% CI 0.43-0.88, *P*<0.01; Table 2). This was attributed to a decline in moderate/late preterm births (aOR 0.53, 95% CI 0.35-0.80, *P*<0.01; Table 2). To confirm that this was a consistent finding compared with each preceding year, 2020 was set as the reference year and the adjusted odds of moderate/late preterm birth was ∼1.6-2.4-times higher in all preceding years (Supplementary Table 1). There were no differences in extremely preterm or very preterm births between year 2020 and the preceding seven years (Table 2, Supplementary Table 1).

**Table 2.** Distribution of singleton livebirths by gestational age category for each year, and odds of preterm singleton livebirths in year 2020 compared with consolidated 2013-2019 data.

|  | 2013 | 2014 | 2015 | 2016 | 2017 | 2018 | 2019 | 2020 | 2013-2019 | 2020 | 2020 vs consolidated 2013-2019 |  |
| --- | --- | --- | --- | --- | --- | --- | --- | --- | --- | --- | --- | --- |
|  | N (%) | N (%) | N (%) | N (%) | N (%) | N (%) | N (%) | N (%) | Per 1,000 (SD) | Per 1,000 | uOR (95% CI) | aOR (95% CI) |
| *Early restriction period* |  |  |  |  |  |  |  |  |  |  |  |  |
| *16 March – 17 April* |  |  |  |  |  |  |  |  |  |  |  |  |
| **Total preterm** | 64 (7.7) | 80 (9.1) | 59 (6.7) | 68 (7.8) | 61 (7.0) | 66 (7.6) | 71 (7.8) | 41 (4.9) | 76.67 (0.007) | 48.93 | 0.62 (0.45-0.86)* | 0.62 (0.43-0.88)* |
| **Full-term** | 765 (92.3) | 802 (90.9) | 818 (93.3) | 807 (92.2) | 807 (93.0) | 806 (92.4) | 843 (92.2) | 797 (95.1) | 923.33 (0.007) | 951.07 | - | - |
| **Extremely preterm** | 6 (0.7) | 8 (0.9) | 5 (0.6) | 5 (0.6) | 4 (0.5) | 5 (0.6) | 8 (0.9) | 7 (0.8) | 6.70 (0.002) | 8.35 | 1.21 (0.54-2.71) | 1.70 (0.69-4.20) |
| **Very preterm** | 9 (1.1) | 9 (1.0) | 10 (1.1) | 8 (0.9) | 11 (1.3) | 8 (0.9) | 9 (1.0) | 5 (0.6) | 10.46 (0.001) | 5.97 | 0.55 (0.22-1.38) | 0.62 (0.22-1.75) |
| **Mod/late preterm** | 49 (5.9) | 63 (7.1) | 44 (5.0) | 55 (6.3) | 46 (5.3) | 53 (6.1) | 54 (5.9) | 29 (3.5) | 59.51 (0.007) | 34.61 | 0.57 (0.38-0.83)* | 0.53 (0.35-0.80)* |
| **Total births** | 829 (100) | 882 (100) | 877 (100) | 875 (100) | 868 (100) | 872 (100) | 914 (100) | 838 (100) | 1000 | 1000 |  |  |
| *Late restriction period* |  |  |  |  |  |  |  |  |  |  |  |  |
| *30 March – 1 May* |  |  |  |  |  |  |  |  |  |  |  |  |
| **Total preterm** | 61 (7.5) | 79 (8.9) | 60 (6.8) | 68 (7.8) | 61 (7.1) | 51 (6.1) | 68 (7.5) | 53 (5.9) | 73.93 (0.009) | 59.35 | 0.79 (0.59-1.06) | 0.74 (0.54-1.03)^ |
| **Full-term** | 751 (92.5) | 805 (91.1) | 826 (93.2) | 806 (92.2) | 803 (92.9) | 788 (93.9) | 833 (92.5) | 840 (94.1) | 926.07 (0.009) | 940.65 | - | - |
| **Extremely preterm** | 6 (0.7) | 4 (0.5) | 3 (0.3) | 6 (0.7) | 6 (0.7) | 5 (0.6) | 7 (0.8) | 5 (0.6) | 6.11 (0.002) | 5.60 | 0.90 (0.35-2.30) | 1.02 (0.35-2.94) |
| **Very preterm** | 8 (1.0) | 11 (1.2) | 8 (0.9) | 7 (0.8) | 7 (0.8) | 7 (0.8) | 10 (1.1) | 6 (0.7) | 9.57 (0.002) | 6.72 | 0.69 (0.30-1.61) | 0.63 (0.22-1.77) |
| **Mod/late preterm** | 47 (5.8) | 64 (7.2) | 49 (5.5) | 55 (6.3) | 48 (5.6) | 39 (4.6) | 51 (5.7) | 42 (4.7) | 58.25 (0.008) | 47.03 | 0.80 (0.57-1.10) | 0.73 (0.51-1.05)^ |
| **Total births** | 812 (100) | 884 (100) | 886 (100) | 874 (100) | 864 (100) | 839 (100) | 901 (100) | 893 (100) | 1000 | 1000 |  |  |

uOR: unadjusted odds ratio; aOR: adjusted odds ratio for maternal age, body mass index, ethnicity, parity, socioeconomic status by residence, and history or current asthma, diabetes, and/or hypertensive disorder. \* $P < 0.01$ . ^ $P = 0.073-0.087$ .

During the “late” period in 2020, the prevalence of preterm birth was 59 per 1,000 singleton births versus an average of 74 during the same calendar weeks of the previous seven years (aOR 0.74, 95% CI 0.54-1.03, *P*=0.07; Table 2). This was attributed to a trending reduction in moderate/late preterm births in the adjusted model (aOR 0.73, 95% CI 0.51-1.05, *P*=0.09; Table 2, Supplementary Table 1).

We then separated the analyses by type of birth onset. A reduction in planned, but not spontaneous, moderate/late preterm births was seen during the “early” period compared with consolidated 2013-2019 data (aOR 0.39, 95% CI 0.22-0.71, *P*<0.01; Table 3). When 2020 was set as the reference year, the adjusted odds of planned moderate/late preterm birth during the “early” period was 2.2-3.3-times greater in all preceding years (Supplementary Table 2). The reduction in planned moderate/late preterm birth was less pronounced during the “late” period (aOR 0.61, 95% CI 0.38-0.99, *P*<0.05; Table 3, Supplementary Table 2).

**Table 3.** Odds of preterm singleton livebirths by birth onset (planned or spontaneous) in year 2020 compared with consolidated 2013-2019 data.

|  | Planned births |  |  |  | Spontaneous births |  |  |  |
| --- | --- | --- | --- | --- | --- | --- | --- | --- |
|  | 2013-2019 | 2020 | 2020 vs consolidated 2013-2019 |  | 2013-2019 | 2020 | 2020 vs consolidated 2013-2019 |  |
|  | Per 1,000 (SD) | Per 1,000 | uOR (95% CI) | aOR (95% CI) | Per 1,000 (SD) | Per 1,000 | uOR (95% CI) | aOR (95% CI) |
| *Early restriction period* |  |  |  |  |  |  |  |  |
| *16 March – 17 April* |  |  |  |  |  |  |  |  |
| **Total preterm** | 80.37 (0.011) | 44.06 | 0.53 (0.34-0.82)* | 0.50 (0.31-0.83)* | 72.06 (0.012) | 57.14 | 0.78 (0.48-1.28) | 0.80 (0.48-1.36) |
| **Full-term** | 919.63 (0.011) | 955.94 | - | - | 927.94 (0.012) | 942.86 | - | - |
| **Extremely preterm** | 5.30 (0.003) | 7.66 | 1.39 (0.47-4.13) | 2.31 (0.61-8.66) | 8.46 (0.003) | 9.52 | 1.11 (0.33-3.71) | 1.33 (0.37-4.75) |
| **Very preterm** | 11.48 (0.002) | 7.66 | 0.64 (0.23-1.81) | 0.72 (0.21-2.44) | 9.19 (0.003) | 3.18 | 0.34 (0.05-2.52) | 0.38 (0.05-2.98) |
| **Mod/late preterm** | 63.59 (0.009) | 28.74 | 0.44 (0.26-0.74)* | 0.39 (0.22-0.71)* | 54.41 (0.013) | 44.44 | 0.80 (0.46-1.41) | 0.77 (0.43-1.40) |
| *Late restriction period* |  |  |  |  |  |  |  |  |
| *30 March – 1 May* |  |  |  |  |  |  |  |  |
| **Total preterm** | 75.35 (0.018) | 51.60 | 0.67 (0.45-0.99)* | 0.66 (0.42-1.01)^ | 72.15 (0.016) | 72.73 | 1.01 (0.65-1.57) | 0.93 (0.57-1.51) |
| **Full-term** | 924.65 (0.034) | 948.40 | - | - | 927.85 (0.016) | 927.27 | - | - |
| **Extremely preterm** | 3.25 (0.002) | 5.34 | 1.60 (0.45-5.76) | 1.48 (0.31-7.10) | 9.72 (0.005) | 6.06 | 0.62 (0.15-2.64) | 0.83 (0.19-3.60) |
| **Very preterm** | 9.75 (0.024) | 5.34 | 0.53 (0.16-1.75) | 0.70 (0.21-2.40) | 9.35 (0.003) | 9.09 | 0.97 (0.29-3.24) | 0.49 (0.06-3.73) |
| **Mod/late preterm** | 62.35 (0.019) | 40.92 | 0.64 (0.41-0.99)* | 0.61 (0.38-0.99)* | 53.08 (0.015) | 57.58 | 1.09 (0.66-1.78) | 0.99 (0.58-1.68) |

uOR: unadjusted odds ratio; aOR: adjusted odds ratio for maternal age, body mass index, ethnicity, parity, socioeconomic status by residence, and history or current asthma, diabetes, and/or hypertensive disorder. \* $P < 0.01$ . ^ $P = 0.057$ .

To capture the two-week spike in spontaneous preterm births in 2020 (26 April to 9 May), a logistic regression was performed. Compared with 2020, the odds of preterm versus full-term spontaneous birth was lower only in 2014 (aOR 0.50, 95% CI 0.24-1.07, *P*=0.07) and 2017 (aOR 0.49, 95% CI 0.22-1.10, *P*=0.08, Supplementary Table 3).

Between March 16 to May 1, the rate of stillbirth for all singleton pregnancies (19-43 weeks’ gestation) did not differ between the years (*X*^2^(7, *N*=10,044) = 4.680, *P*=0.70; year: *N* (%): 2013: 8 (0.7); 2014: 5 (0.4); 2015: 4 (0.3); 2016: 8 (0.6); 2017: 8 (0.6); 2018: 4 (0.3); 2019: 4 (0.3); 2020: 6 (0.5)). During the two-spike in spontaneous preterm births in 2020 (26 April to 9 May), singleton stillbirth rates did not differ between the years (*X*^2^(7, *N*=3,055) = 2.758, *P*=0.91; year: *N* (%): 2013: 4 (1.1); 2014: 2 (0.5); 2015: 4 (1.0); 2016: 3 (0.8); 2017: 3 (0.8); 2018: 2 (0.5); 2019: 3 (0.8); 2020: 1 (0.3)). There were no singleton neonatal deaths reported between 26 April to 9 May, 2020.

For multiple pregnancies, where at least one infant in the set was born live, the odds of preterm birth did not differ in year 2020 compared with the preceding seven years (Supplementary Table 4).

## Discussion

In this study, at a tertiary perinatal hospital in Queensland, Australia, a significant reduction in preterm births was observed following the implementation of measures to contain the spread of COVID-19. The reduction was driven primarily by a decline in moderate/late preterm infants, and the greatest impact was seen during the earliest period of restrictions. Furthermore, this reduction appeared to be attributed to planned, but not spontaneous, preterm births. These findings pertain to singleton, and not multiple, pregnancies. Our data contribute to the growing evidence from other countries (5-9) and, together, may reveal novel factors linked to preterm birth.

During early restrictions, planned births for moderate/late preterm infants reduced by more than half when compared with the preceding seven years. Given that infection is an indication for planned preterm birth (10), it is possible that reduced physical contact and increased hygiene contributed to our findings. Furthermore, self-isolation may have resulted in reduced work- and social-related stress, improved sleep quality and/or diet, with an overall improvement to pregnancy health, such as controlled blood pressure, and reduced requirement for a planned preterm birth. Other reasons may include more tele-health antenatal appointments and a possible reduction in care-seeking behaviour by pregnant women. Furthermore, given the uncertainty of the situation at the time, and the close locality of the maternity hospital to the general adult’s hospital, it is possible that women avoided seeking care for antenatal concerns that would normally be grounds for a planned preterm birth. Importantly, we did not observe an increase in stillbirth rates during this time.

Almost immediately following the nadir in planned preterm births, there was a spike in spontaneous preterm births from late-April to early-May, although this was largely not significant when compared with previous years. It is possible that the same women who were not captured for a medically indicated planned preterm birth in the preceding two weeks, subsequently went into spontaneous preterm labour. Further analyses of preterm birth rates by gestational weeks using a larger dataset may support this observation. No neonatal deaths were reported during the rise in spontaneous preterm births.

The earliest period of restrictions, before the government-imposed hard lockdown, had the greatest influence on reducing planned preterm birth rates. In Australia, there was a remarkable spike in the “panic index” in early March (11). This did not coincide with any major restrictions to movement or travel, nor local COVID-19 cases, but was likely related to observations by the general population of the international impacts of COVID-19. At this time, hand sanitisers and antibacterial handwashes sold out in most parts of the country, highlighting the magnitude of behaviour change. This timing is somewhat consistent with a large nationwide study from the Netherlands, which reported reductions in moderate/late preterm births in the 2-4-month period following initial mitigation measures, but not when stricter measures were introduced 1-2 weeks later, albeit there was no separation by planned and spontaneous preterm births (7).

We did not see a persistent level of reduction in moderate/late planned preterm births later in the pandemic, following the implementation of more formal lockdown measures. Given that COVID-19 cases did not rise as initially expected in Australia, it is possible that attending hospital for antenatal concerns in these later weeks was no longer avoided. Alternatively, unintended consequences of prolonged isolation, such as reduced physical activity and mental health concerns, may have counterbalanced the benefits from other, early behavioural changes that impact on pregnancy health. A study from Canada recruited pregnant and new mothers between mid-April and early May and found that self-reported measures of reduced physical activity were reduced, and levels of depression and anxiety were increased compared with pre-pandemic levels (12). During the hard lockdown in Australia specifically, mental health concerns including depression and anxiety were widespread in the general population (13).

A nationwide study in Denmark found no effect on moderate/late or very preterm births during the strictest month of lockdown compared with the same calendar period in the previous five years (6). However, a significant reduction in extremely preterm births was observed. Similarly, in a designated area of Ireland, the proportion of extremely-and very-low-birth-weight infants was unusually low in the first four months of 2020 compared with the same period in the preceding 19 years (8). It is difficult to ascertain why the Danish and Irish observed reductions in the earliest and smallest infants (6, 8) while we, the Dutch (7), and Italians (5) observed reductions in moderate/late preterm births only. Furthermore, there have been reports of adverse neonatal outcomes during lockdown, including a 1.5-fold increase in stillbirth and 3-fold increase in neonatal mortality in Nepal (4), a 6-fold increase in stillbirth in a London hospital (3), and a 2.6-fold increase in stillbirth in a large region in Italy (5). The implementation of formal COVID-19 restriction measures and population responses have varied across the globe (11, 14), which may underlie some of the observed differences.

### Limitations

This was a retrospective data collection approach using a hospital database that had some missing data. We included all livebirths ≥23 completed weeks’ gestation, as active resuscitation and support is usually offered from this gestation depending on parental wishes. While our statistical analyses also included some babies who later died in hospital during March 16-May 1, 2013-2020 (*N*=21/9,986 singleton livebirths), we feel this is a better representation of the birthing population, especially of those born preterm. While there were no neonatal deaths during the spike in spontaneous preterm births, we did not examine other variables related to neonatal health (e.g. Apgar scores). It is also possible that we received fewer referrals from other local perinatal centres during the restriction period, due to patient avoidance of the tertiary setting.

## Conclusions

Preterm birth is the leading cause of neonatal death globally, and those who survive are at greater risk of cognitive, behavioural, motor, and respiratory impairments (15-18). The preterm births rate is increasing in most parts of the world, mainly attributed to increases in planned preterm births (10, 19, 20). We show that our early response to the COVID-19 pandemic was associated with an unprecedented reduction in planned moderate/late preterm births. This aligns with studies from other countries (5-9), albeit differences are seen with respect to which category of preterm infants were most positively affected and we are the first to differentiate between planned and spontaneous births. A global effort is now exploring the links between COVID-19 restrictions, preterm births, regional variation, and temporal trends (https://www.ipopstudy.com/).

## Data Availability

Data may be made available upon request

## Acknowledgements

The authors wish to acknowledge the contributions of Yanlin Liu for data extraction through Mater Data and Analytics, and Nicholas Matigian for statistical support through the Research and Statistical Support Service, Faculty of Medicine, provided by QCIF Facility for Advanced Bioinformatics (QFAB), The University of Queensland.

---

**Supplementary Table 1.**
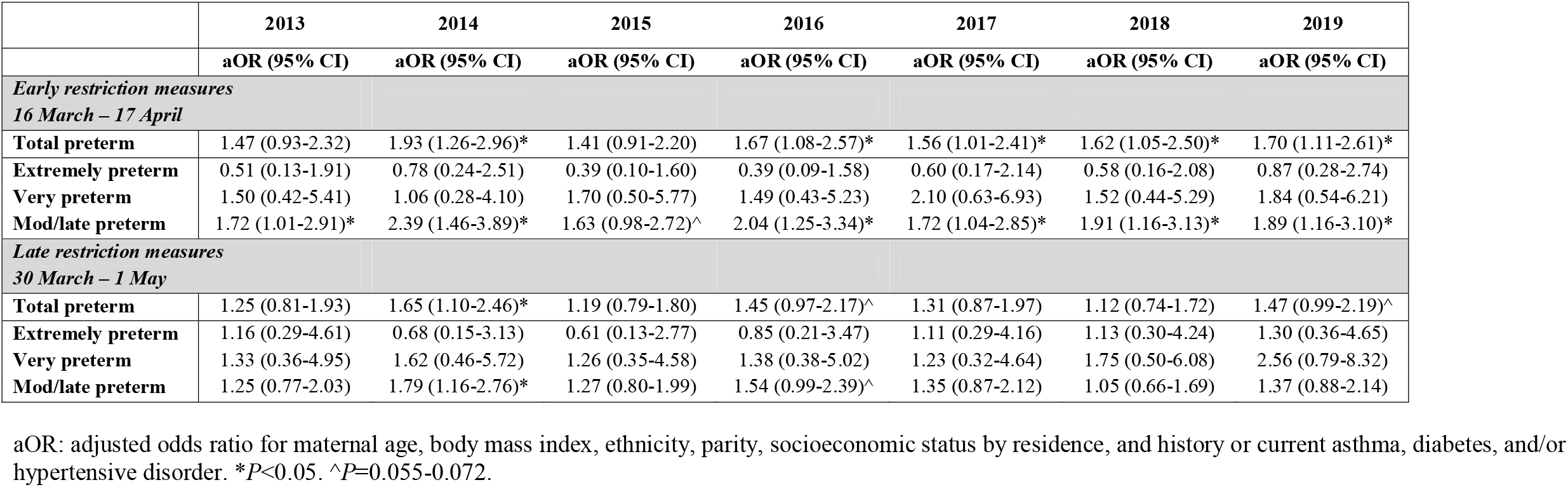
Odds of preterm singleton livebirths in each year compared with year 2020.

|  | 2013 | 2014 | 2015 | 2016 | 2017 | 2018 | 2019 |
| --- | --- | --- | --- | --- | --- | --- | --- |
|  | aOR (95% CI) | aOR (95% CI) | aOR (95% CI) | aOR (95% CI) | aOR (95% CI) | aOR (95% CI) | aOR (95% CI) |
| *Early restriction measures* |  |  |  |  |  |  |  |
| *16 March – 17 April* |  |  |  |  |  |  |  |
| **Total preterm** | 1.47 (0.93-2.32) | 1.93 (1.26-2.96)* | 1.41 (0.91-2.20) | 1.67 (1.08-2.57)* | 1.56 (1.01-2.41)* | 1.62 (1.05-2.50)* | 1.70 (1.11-2.61)* |
| **Extremely preterm** | 0.51 (0.13-1.91) | 0.78 (0.24-2.51) | 0.39 (0.10-1.60) | 0.39 (0.09-1.58) | 0.60 (0.17-2.14) | 0.58 (0.16-2.08) | 0.87 (0.28-2.74) |
| **Very preterm** | 1.50 (0.42-5.41) | 1.06 (0.28-4.10) | 1.70 (0.50-5.77) | 1.49 (0.43-5.23) | 2.10 (0.63-6.93) | 1.52 (0.44-5.29) | 1.84 (0.54-6.21) |
| **Mod/late preterm** | 1.72 (1.01-2.91)* | 2.39 (1.46-3.89)* | 1.63 (0.98-2.72)^ | 2.04 (1.25-3.34)* | 1.72 (1.04-2.85)* | 1.91 (1.16-3.13)* | 1.89 (1.16-3.10)* |
| *Late restriction measures* |  |  |  |  |  |  |  |
| *30 March – 1 May* |  |  |  |  |  |  |  |
| **Total preterm** | 1.25 (0.81-1.93) | 1.65 (1.10-2.46)* | 1.19 (0.79-1.80) | 1.45 (0.97-2.17)^ | 1.31 (0.87-1.97) | 1.12 (0.74-1.72) | 1.47 (0.99-2.19)^ |
| **Extremely preterm** | 1.16 (0.29-4.61) | 0.68 (0.15-3.13) | 0.61 (0.13-2.77) | 0.85 (0.21-3.47) | 1.11 (0.29-4.16) | 1.13 (0.30-4.24) | 1.30 (0.36-4.65) |
| **Very preterm** | 1.33 (0.36-4.95) | 1.62 (0.46-5.72) | 1.26 (0.35-4.58) | 1.38 (0.38-5.02) | 1.23 (0.32-4.64) | 1.75 (0.50-6.08) | 2.56 (0.79-8.32) |
| **Mod/late preterm** | 1.25 (0.77-2.03) | 1.79 (1.16-2.76)* | 1.27 (0.80-1.99) | 1.54 (0.99-2.39)^ | 1.35 (0.87-2.12) | 1.05 (0.66-1.69) | 1.37 (0.88-2.14) |

aOR: adjusted odds ratio for maternal age, body mass index, ethnicity, parity, socioeconomic status by residence, and history or current asthma, diabetes, and/or hypertensive disorder. \* $P < 0.05$ . ^ $P = 0.055-0.072$ .

**Supplementary Table 2.**
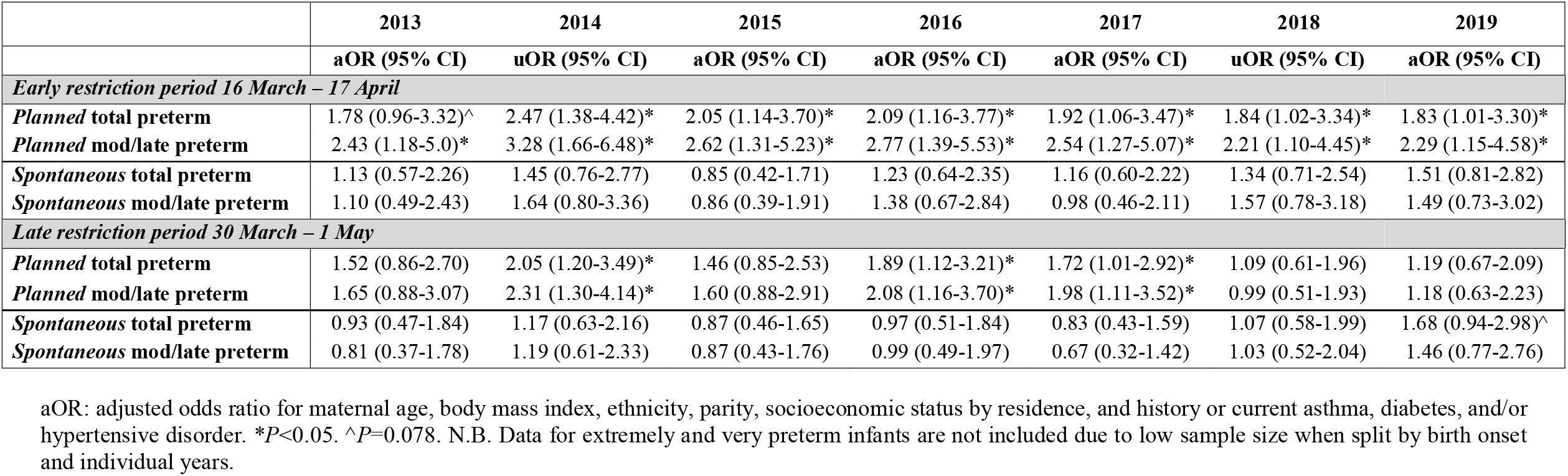
Odds of preterm singleton livebirths by birth onset (planned or spontaneous) in each year compared with year 2020.

|  | 2013 | 2014 | 2015 | 2016 | 2017 | 2018 | 2019 |
| --- | --- | --- | --- | --- | --- | --- | --- |
|  | aOR (95% CI) | uOR (95% CI) | aOR (95% CI) | aOR (95% CI) | aOR (95% CI) | uOR (95% CI) | aOR (95% CI) |
| *Early restriction period 16 March – 17 April* |  |  |  |  |  |  |  |
| **Planned total preterm** | 1.78 (0.96-3.32)^ | 2.47 (1.38-4.42)* | 2.05 (1.14-3.70)* | 2.09 (1.16-3.77)* | 1.92 (1.06-3.47)* | 1.84 (1.02-3.34)* | 1.83 (1.01-3.30)* |
| **Planned mod/late preterm** | 2.43 (1.18-5.0)* | 3.28 (1.66-6.48)* | 2.62 (1.31-5.23)* | 2.77 (1.39-5.53)* | 2.54 (1.27-5.07)* | 2.21 (1.10-4.45)* | 2.29 (1.15-4.58)* |
| **Spontaneous total preterm** | 1.13 (0.57-2.26) | 1.45 (0.76-2.77) | 0.85 (0.42-1.71) | 1.23 (0.64-2.35) | 1.16 (0.60-2.22) | 1.34 (0.71-2.54) | 1.51 (0.81-2.82) |
| **Spontaneous mod/late preterm** | 1.10 (0.49-2.43) | 1.64 (0.80-3.36) | 0.86 (0.39-1.91) | 1.38 (0.67-2.84) | 0.98 (0.46-2.11) | 1.57 (0.78-3.18) | 1.49 (0.73-3.02) |
| *Late restriction period 30 March – 1 May* |  |  |  |  |  |  |  |
| **Planned total preterm** | 1.52 (0.86-2.70) | 2.05 (1.20-3.49)* | 1.46 (0.85-2.53) | 1.89 (1.12-3.21)* | 1.72 (1.01-2.92)* | 1.09 (0.61-1.96) | 1.19 (0.67-2.09) |
| **Planned mod/late preterm** | 1.65 (0.88-3.07) | 2.31 (1.30-4.14)* | 1.60 (0.88-2.91) | 2.08 (1.16-3.70)* | 1.98 (1.11-3.52)* | 0.99 (0.51-1.93) | 1.18 (0.63-2.23) |
| **Spontaneous total preterm** | 0.93 (0.47-1.84) | 1.17 (0.63-2.16) | 0.87 (0.46-1.65) | 0.97 (0.51-1.84) | 0.83 (0.43-1.59) | 1.07 (0.58-1.99) | 1.68 (0.94-2.98)^ |
| **Spontaneous mod/late preterm** | 0.81 (0.37-1.78) | 1.19 (0.61-2.33) | 0.87 (0.43-1.76) | 0.99 (0.49-1.97) | 0.67 (0.32-1.42) | 1.03 (0.52-2.04) | 1.46 (0.77-2.76) |

aOR: adjusted odds ratio for maternal age, body mass index, ethnicity, parity, socioeconomic status by residence, and history or current asthma, diabetes, and/or hypertensive disorder. \* $P < 0.05$ . ^ $P = 0.078$ . N.B. Data for extremely and very preterm infants are not included due to low sample size when split by birth onset and individual years.

**Supplementary Table 3.** Odds of preterm singleton livebirths by birth onset (planned or spontaneous) in each year compared with year 2020 easing restrictions/post-lockdown period.

|  | 2013 | 2014 | 2015 | 2016 | 2017 | 2018 | 2019 | 2020 |
| --- | --- | --- | --- | --- | --- | --- | --- | --- |
| *Easing restrictions/post-lockdown period 26 April – 9 May* |  |  |  |  |  |  |  |  |
| **Total preterm [N, (%)]** | 34 (90.5) | 30 (7.7) | 26 (6.5) | 33 (8.3) | 25 (7.0) | 21 (5.7) | 30 (7.6) | 32 (8.5) |
| **Full-term [N, (%)]** | 323 (9.5) | 359 (92.3) | 371 (93.5) | 363 (91.7) | 331 (93.0) | 347 (94.3) | 363 (92.4) | 344 (91.5) |
| **Total [N, (%)]** | 357 (100) | 389 (100) | 397 (100) | 396 (100) | 356 (100) | 368 (100) | 393 (100) | 376 (100) |
| *Planned preterm* |  |  |  |  |  |  |  |  |
| **uOR (95% CI) vs. 2020** | 2.48 (1.14-5.42)* | 1.58 (0.68-3.63) | 0.85 (0.34-2.13) | 1.87 (0.85-4.12) | 1.60 (0.70-3.65) | 0.88 (0.35-2.22) | 1.23 (0.53-2.86) | - |
| **aOR (95% CI) vs. 2020** | 2.24 (0.90-5.57)^ | 1.83 (0.72-4.65) | 0.76 (0.27-2.08) | 2.12 (0.88-5.11) | 1.90 (0.77-4.70) | 0.894 (0.35-2.54) | 1.40 (0.55-3.52) | - |
| *Spontaneous preterm* |  |  |  |  |  |  |  |  |
| **uOR (95% CI) vs. 2020** | 0.55 (0.27-1.13) | 0.57 (0.29-1.13) | 0.70 (0.36-1.38) | 0.57 (0.28-1.16) | 0.46 (0.21-1.00)* | 0.55 (0.26-1.15) | 0.74 (0.37-1.44) | - |
| **aOR (95% CI) vs. 2020** | 0.65 (0.30-1.42) | 0.50 (0.24-1.07)^ | 0.77 (0.38-1.57) | 0.61 (0.30-1.28) | 0.49 (0.22-1.10)^ | 0.58 (0.27-1.24) | 0.78 (0.39-1.56) | - |

aOR: adjusted odds ratio for maternal age, parity, socioeconomic status by residence, and history or current diabetes and/or hypertensive disorder. \* $P < 0.05$ . ^ $P = 0.073$ - $0.083$ . N.B. Data not presented by gestational age category due to low sample size when split by birth onset and individual years for this two-week period.

**Supplementary Table 4.** Distribution and odds of preterm multiple livebirths between 16 March-1 May each year compared with 2020.

|  | 2013 | 2014 | 2015 | 2016 | 2017 | 2018 | 2019 | 2020 |
| --- | --- | --- | --- | --- | --- | --- | --- | --- |
| **Preterm [N, (%)]** | 25 (67.6) | 22 (73.3) | 23 (67.6) | 22 (78.6) | 21 (80.8) | 18 (75.0) | 16 (64.0) | 15 (75.0) |
| **Full-term [N, (%)]** | 12 (32.4) | 8 (26.7) | 11 (32.4) | 6 (21.4) | 5 (19.2) | 6 (25.0) | 9 (36.0) | 5 (25.0) |
| **Total [N, (%)]** | 37 (100)<sup>a</sup> | 30 (100)<sup>b</sup> | 34 (100)<sup>b</sup> | 28 (100)<sup>b</sup> | 26 (100) | 24 (100) | 25 (100) | 20 (100) |
| **Preterm uOR (95% CI) vs. 2020** | 1.44 (0.42-4.90) | 1.09 (0.30-3.99) | 1.44 (0.42-4.96) | 0.82 (0.21-3.18) | 0.71 (0.18-2.91) | 1.00 (0.25-3.94) | 1.69 (0.46-6.20) | - |
| **Preterm aOR (95% CI) vs. 2020** | 1.66 (0.46-5.97) | 1.19 (0.31-4.58) | 1.37 (0.38-4.98) | 0.68 (0.17-2.71) | 0.56 (0.13-2.48) | 1.24 (0.30-5.16) | 1.73 (0.45-6.62) | - |

<sup>a</sup> includes two sets of triplets; <sup>b</sup> includes one set of triplets. uOR: unadjusted odds ratio; aOR: Adjusted odds ratio for maternal ethnicity and socioeconomic status by residence.

